# Three-Dimensional virtual reality-based visualization of fetal cardiac anatomy using spatio-temporal image correlation (STIC) ultrasound datasets of normal and abnormal hearts

**DOI:** 10.1101/2024.09.11.24313355

**Authors:** Balu Vaidyanathan, Harikrishnan Anil Maya, Sarin Xavier, Mahesh Kappanayil

## Abstract

This case report demonstrates the feasibility of creating immersive 3D visualizations (3D Virtual reality and 3D printing) from fetal echocardiographic volume datasets for both normal heart and a heart with transposition of great arteries. Immersive 3D technologies could emerge as powerful tools in future for understanding fetal cardiac anatomy for clinical decision making as well as training and research.

## Case Report

Conventional imaging of the fetal heart is done using standard 2-dimensional planes obtaining multiple cross-sectional images of the cardiac structures (1). Newer techniques like 3-dimensional spatio-temporal image correlation (STIC) permits acquisition of volume datasets, followed by multi-planar reconstruction and display of the rendered images (2). Cardiac magnetic resonance imaging has been used in the evaluation of the fetal heart especially in the visualization of the great arteries and extra-cardiac vascular structures (3,4). However, these techniques fail to provide a holistic spatial visualization of the cardiac structures with a limited ability to manipulate the datasets in multiple planes. New developments in 3D technologies - Extended Reality and 3D printing – provide powerful means of immersive visualization (5). Virtual Reality (VR) presents volumetric scan data as digital twins in deeply immersive computer-generated 3D environments using specialized software platforms and hardware (headsets/eyewear) (6). Digital tools provide powerful, means of interaction and engagement with the virtual anatomical 3D models. While CT/MRI scans can be rendered in high quality VR with relative ease, rendering cardiac ultrasound data has been a challenge owing to lower spatial resolution and unfavorable signal-noise ratios (7). Challenges in converting raw data to formats amenable for processing by 3D modelling software and difficulties in segmentation have been among technical barriers to VR visualization of fetal hearts (6,8). This report explores technical feasibility of 3D VR visualization and 3D printing of fetal cardiac ultrasound data of normal and abnormal hearts acquired using STIC technology.

The study was conducted in a tertiary pediatric cardiac facility of a university hospital in Kerala, South India. A dedicated fetal cardiology referral service was established in 2008 and was escalated into a dedicated division in 2016. The center has an advanced Point-of-Care Medical 3D printing and Extended Reality (XR) laboratory for cardiovascular care.

The fetal heart datasets were recorded using Voluson Expert 22 equipment (GE healthcare, Zipf, Austria) using volume (RM7C) and electronic matrix array (eM6C, 3^rd^ generation) probes using spatio-temporal image correlation (STIC) settings. In 2D and 3D settings, speckle reduction imaging (SRI) and compound resolution (crossXbeam) imaging (CRI) settings were optimized in order to acquire volume datasets having a frame rate of 250-300 Hz after acquisition. After acquisition, the datasets were exported in cartesian.vol format to a VR-capable workstation with Graphic Processing Unit (GPU). The raw ultrasound data was converted to a compatible digital 3D format (NIfTI) using open source 3DSlicer image computing platform (https://www.slicer.org). FDA-approved proprietary medical 3D modelling and surgical planning software Elucis (Elucis Next v1.9.5, Realize Medical, Canada) and Meta Quest2 VR device were used for VR rendering and visualization. Data was first viewed in 2D grey-scale format within the VR environment displaying the regions of interest (ROI). Blood-pools within the heart and both outflows were carefully segmented using thresholding tools across multiple 2D image slices, creating 3D blood-pool models.

This required defining the image-intensity thresholds of the ROI and demarcating the borders across every slice of data, assisted by 3D interpolation tools, finally producing 3D representation of the blood-pool. Creation of 1mm shell around the blood-pool, followed by Boolean subtraction of the latter created ‘hollow-heart’ models. Handheld controllers and digital tools were used to engage with the 3D models in VR. Blood-pool models were exported as. stl (standard triangulation language) files and 3D printed (for added validation) using photopolymeric clear resin on Form 3BL (Formlabs, USA) 3D printer. The typical time required for VR formatting was about an hour while 3D printing needed another 90-120 minutes. Appropriate patient consent was obtained for the study.

We selected 2 fetal heart volume datasets, a 22-week gestation normal heart and a 31-week heart with D-transposition of great arteries. Simultaneous viewing of the 3D casts superimposed upon the grey-scale 2D slices allowed correlation and validation of 3D with 2D anatomy. Spatial relationships between cardiac chambers, normal origins of great arteries from respective ventricles and their crossing were clearly observed in 3D VR of the normal heart (Figures 1A-H, 3A). Parallel course and transposed great artery origins were similarly clearly identified in the fetus with d-TGA (figures 2A-F, 3C). Collaboration tool within the software allowed multiple team members to join VR session in real-time, and actively engage on the same model. Blood-pool 3D models (Figure 3B,D)created in VR were successfully 3D printed in 1:1 scale, providing opportunity for physical interaction with the fetal cardiac anatomy.

**Figure 1.**
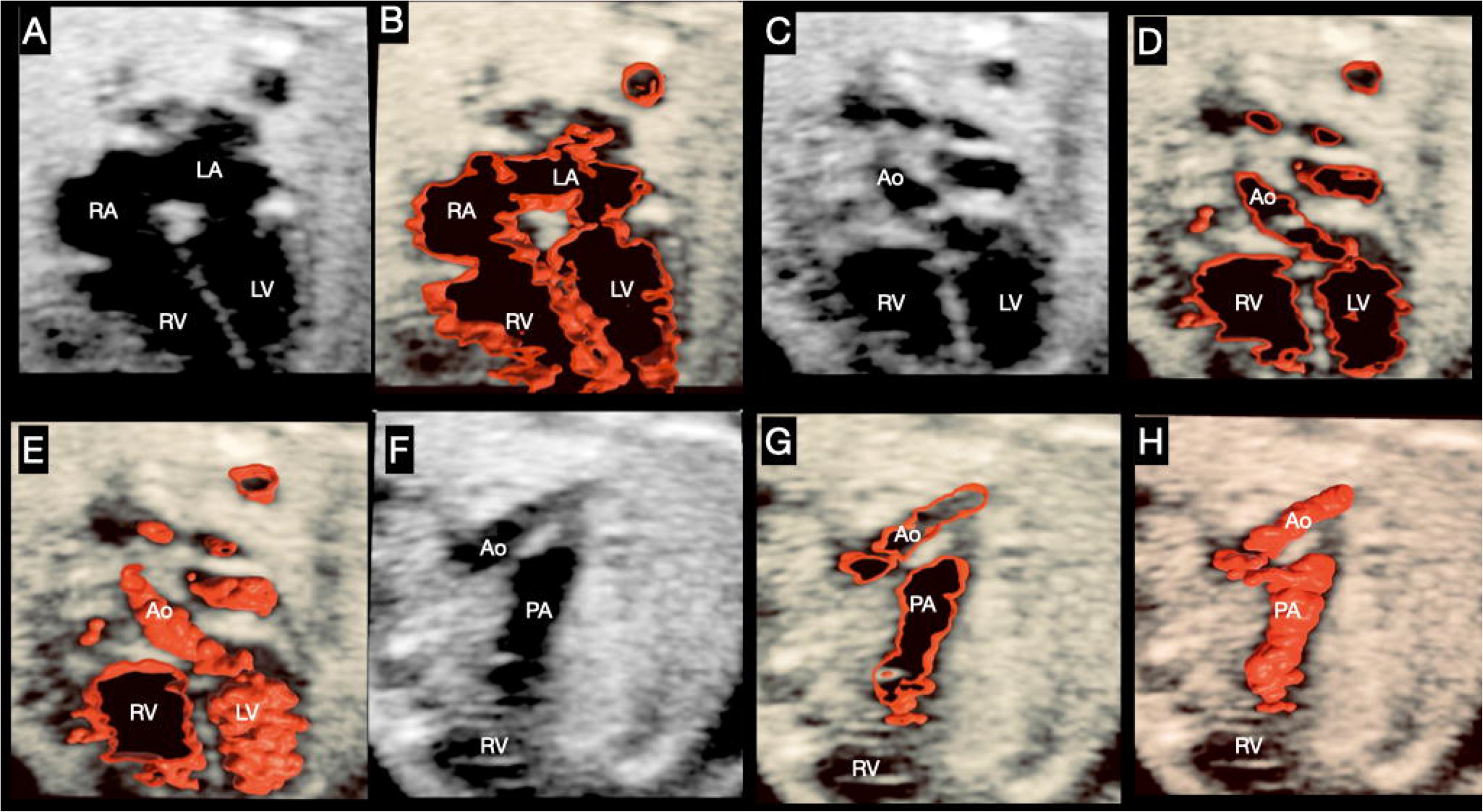
shows the correlation of 2D grey-scale slices of cardiac ultrasound data and 3D models in a case with normal fetal cardiac anatomy, in Virtual Reality using 3D modelling software. 1A shows axial 2D slice depicting 4-chamber view. 1B shows 3D hollow cast digitally created around the blood-pool, superimposed upon the grey-scale slice. 1C shows 2D slice with the left ventricular outflow tract; 1D shows the 3D hollow cast at the corresponding slice level. Figure 1E shows the 3D cast of the LV and LV outflow extending anterior to the plane of the 2D slice. Images 1F, G, H show the RV outflow in axial grey-scale 2D slice, 3D cast at the slice level, and 3D cast extending in a further anterior plane respectively. 3D model clearly shows anatomically appropriate origin of the great arteries and their normal spatial relationship with crossing great arteries. RV – right ventricle, LV-left ventricle, Ao-Aorta, PA-pulmonary artery

**Figure 2.**
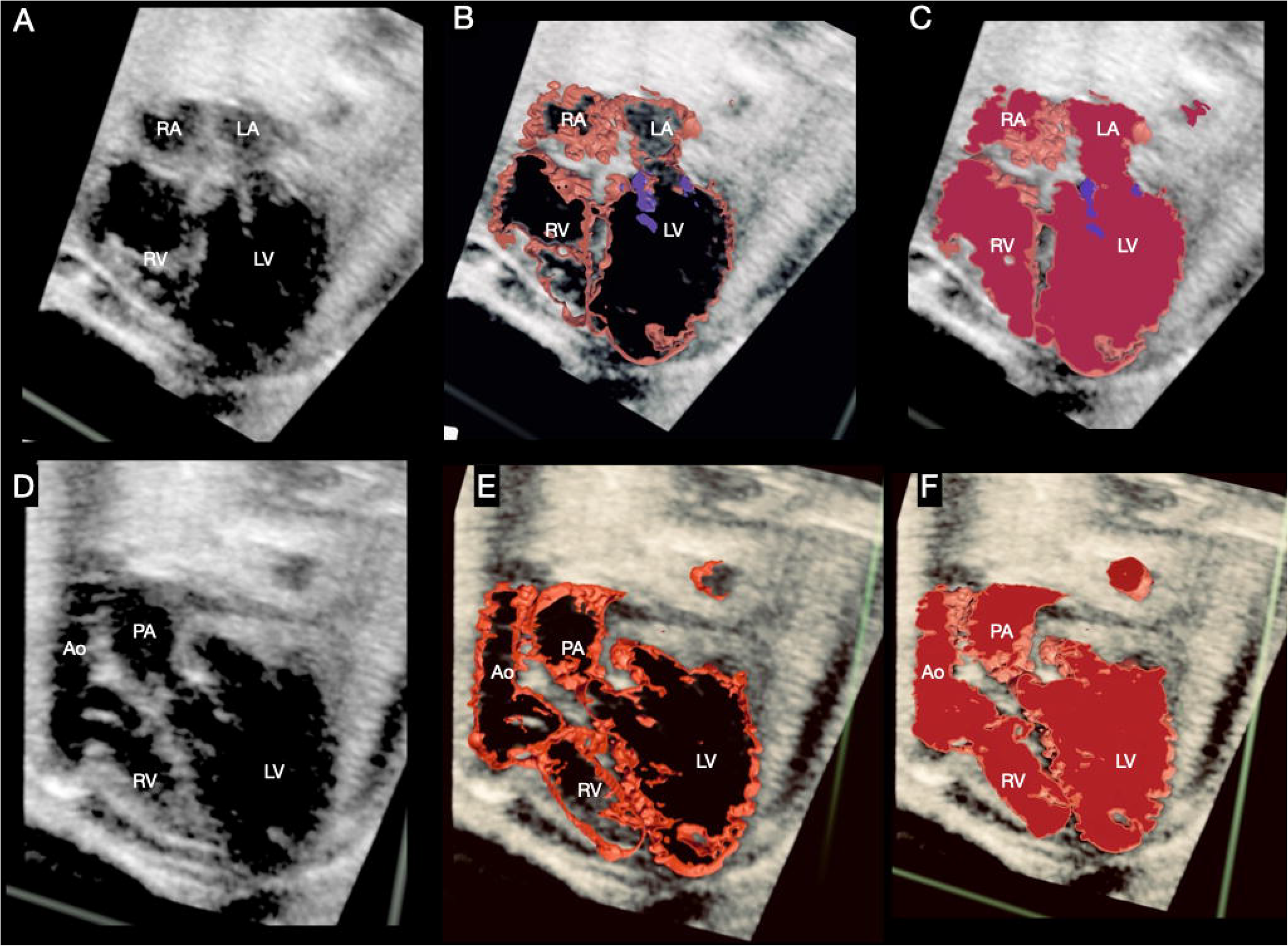
shows VR snapshots of 2D and 3D cast visualization in a case of fetal heart with transposition of great arteries. 2A shows an axial 2D slice of ultrasound data representing 4-chamber view. 2B shows a hollow 3D cast of the blood-pool transected and superimposed on the corresponding 2D slice. 2C shows a solid blood-pool 3D model transected and superimposed on the same corresponding 2D slice. Mitral Valve leaflets have been marked in blue. Satisfactory accuracy of anatomical segmentation can be well appreciated. Figures 2 D, E, F similarly depict ventriculoarterial discordance with parallel great arteries as axial 2D slice(D), 3D hollow cast (E) and 3D solid blood-pool model (F), digitally transected at the same level. RV – right ventricle, LV-left ventricle, Ao-Aorta, PA-pulmonary artery

**Figure 3.**
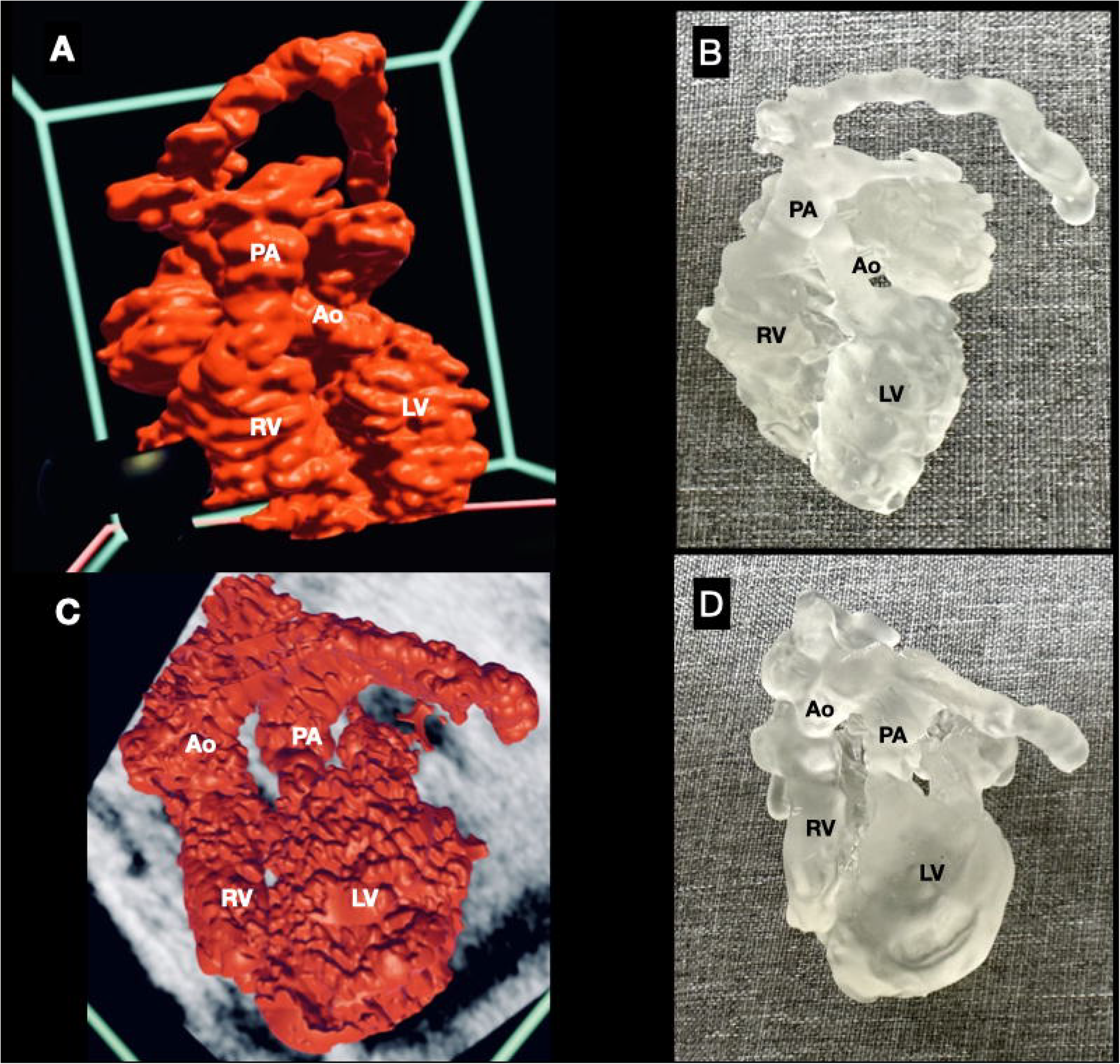
shows 3D models created using VR anatomical modeling. 3A is snapshot of virtual 3D model of case of normal fetal heart as visualized in VR, with crossing great arteries. 3B is a 3D printed prototype of the same file. 3C and D show VR visualization and 3D printed prototype of the 3D model of the fetal heart with D-TGA, showing the parallel great arteries. RV – right ventricle, LV-left ventricle, Ao-Aorta, PA-pulmonary artery

## Discussion

We report the successful utilization of VR technology using fetal heart ultrasound STIC datasets to create immersive 3D VR visualizations and 3D printed models. This was achieved for normal cardiac anatomy as well as for a heart with CHD (transposition of great arteries). VR visualization provided excellent 3D spatial perspective of cardiovascular anatomy, especially the relation of outflow tracts (Figure 3). VR allowed virtual fetal hearts to be grabbed and rotated using handheld controllers, with freedom to view from multiple perspectives(6). ‘Cut-plane’ tool allowed virtual dissection into the hollow-heart, presenting an inside-view into chambers and outflows. VR visualization opens up new opportunities to communicate, teach and collaborate using fetal ultrasound (9). 3D printed models further reinforce the scope and versatility of immersive 3D technologies. Future studies should explore the incremental benefit of this technology for more types of CHD like Double outlet RV and aortic arch anomalies like coarctation of aorta in improving diagnostic accuracy in the prenatal stage. Quality and format of ultrasound data and skills required for segmentation were among the most important challenges in VR conversion (6). Future research and development should focus on addressing these challenges, but also on creating novel dedicated fetal VR software solutions and tools like automatic segmentations and cine-cardiac and flow visualizations.

In conclusion, we demonstrated the feasibility of creating immersive 3D visualizations from fetal echocardiographic volume datasets for both normal and abnormal fetal hearts.Immersive 3D technologies could emerge as powerful tools in future for understanding fetal cardiac anatomy for clinical decision making as well as training and research.

## Data Availability

All data produced in the present study are available upon reasonable request to the authors

## Disclosures

None

## Sources of Funding

None

## Notes

### Competing Interest Statement

The authors have declared no competing interest.

### Funding Statement

This study did not receive any funding

### Author Declarations

This study used electronic health records data ( fetal echocardiography datasets) obtained from human foetuses which were recorded after prior consent from the pregnant woman as per the Pre-conceptional and prenatal diagnostic techniques act of India. The institutional ethics committee of Amrita Institute of Medical Sciences waived the ethical approval for this work since the proper protocol for conduct of fetal echocardiography as per the PcPNT act of India was followed for acquisition of ultrasound datasets and the study involved only use of electronic patient data.

